# Estimating the Asymptomatic Proportion of 2019 Novel Coronavirus onboard the Princess Cruises Ship, 2020

**DOI:** 10.1101/2020.02.20.20025866

**Authors:** Kenji Mizumoto, Katsushi Kagaya, Alexander Zarebski, Gerardo Chowell

**Author notes:** Corresponding author: Kenji Mizumoto, Corresponding author.

## Abstract

The potential infectiousness of asymptomatic COVID-19 cases together with a substantial fraction of asymptomatic infections among all infections, have been highlighted in clinical studies. We conducted statistical modeling analysis to derive the delay-adjusted asymptomatic proportion of the positive COVID-19 infections onboard the Princess Cruises ship along with the timeline of infections. We estimated the asymptomatic proportion at 17.9% (95% CrI: 15.5%–20.2%), with most of the infections occurring before the start of the 2-week quarantine.

## Background

Since COVID-19 emerged in the city of Wuhan, China in December 2019, thousands of people have succumbed to the novel coronavirus especially in the Province of Hubei while hundreds of imported and secondary cases have been reported in multiple countries as of February 29, 2020 [1].

The clinical and epidemiological characteristics of COVID-19 continue to be investigated as the virus continues its march through the human population [2-3]. While reliable estimates of the reproduction number and the death risk associated with COVID-19 are crucially needed to guide public health policy, another key epidemiological parameter that could inform the intensity and range of social distancing strategies to combat COVID-19 is the asymptomatic proportion, which is broadly defined as the proportion of asymptomatic infections among all the infections of the disease. Indeed, the asymptomatic proportion is a useful quantity to gauge the true burden of the disease and better interpret estimates of the transmission potential. This proportion varies widely across infectious diseases, ranging from 8% for measles and 32% for norovirus up to 90-95% for polio [4-6]. Most importantly, it is well established that asymptomatic individuals are frequently able to transmit the virus to others [7-8]. COVID-19 is not the exception to this pattern, with accumulating evidence indicating that a substantial fraction of the infected individuals with the novel coronavirus are asymptomatic [9-11].

As an epidemic progresses over time, suspected cases are examined and tested for the infection using polymerase chain reaction (PCR) or rapid diagnostic test (RDT). Then, time-stamped counts of the test results stratified according to the presence or absence of symptoms at the time of testing are often reported to the public in near real-time. Nevertheless, it is important to note that the estimation of the asymptomatic proportion needs to be handled carefully since real-time outbreak data are influenced by the phenomenon of right censoring, due to the time lag between the time of examination and sample collection and the development of illness.

In this paper, we conduct a statistical modeling analysis to estimate the asymptomatic proportion among infected individuals who have tested positive for COVID-19 infections onboard the Princess Cruises Ship along with their time of infections, accounting for the delay in onset of symptoms and right-censoring.

## Epidemiological description and data

In Yokohama, Japan, an outbreak of COVID-19 unfolded on board the Princess Cruise Ship, which had been under quarantine orders since February 5, 2020, after a former passenger of the cruise ship tested positive for the virus after disembarking in Hong Kong. By February 21, 2020, two days after the scheduled two-week quarantine came to an end, a total of 634 people including one quarantine officer, one nurse and one administrative officer tested positive for COVID-19 out of the 3,711 passengers and crew members on board the vessel. Laboratory testing by PCR were conducted, prioritizing symptomatic or high-risk groups.

Daily time series of laboratory test results for COVID-19 (both positive and negative), including information of presence or absence of symptoms from February 5, 2020 to February 20, 2020 were extracted from secondary sources [12]. The reporting date, number of tests, number of tested positive and number of symptomatic and symptomatic cases at the time of sample collection are provided, while the time of infection and true asymptomatic proportion are not available.

A total of 634 people have tested positive among 3063 tests as of February 20, 2020. Out of 634 cases, a total of 313 cases are female and six were aged 0-19 years, 152 were aged 20-59 years and 476 were 60 years and older (Figure 1). The nationality of the cases includes Japan (270), United States (88), Philippines (54), Canada (51), Australia (49), Hong Kong (30) and China (28).

**Figure 1.**
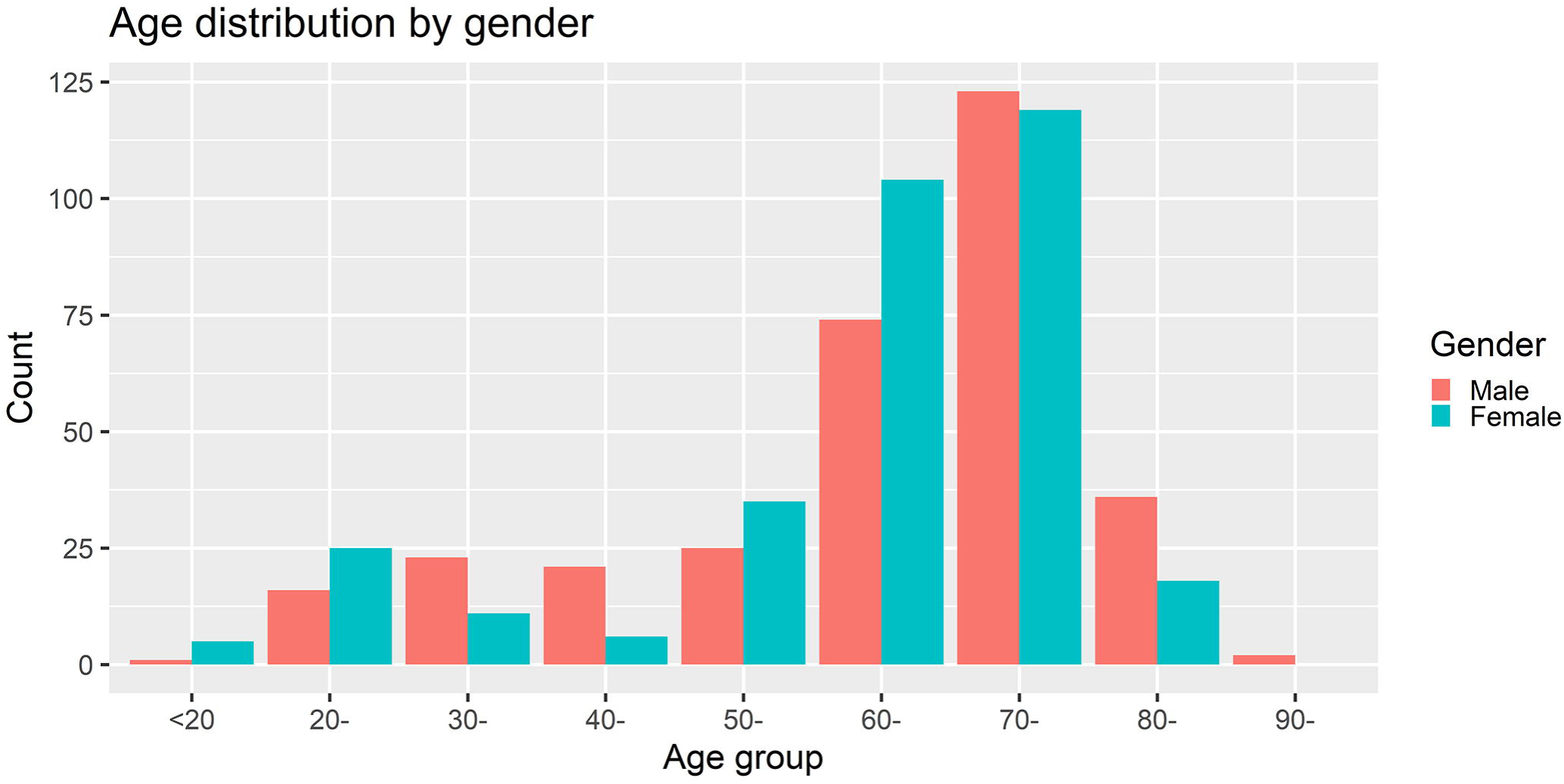
Age distribution of reported 2019 Novel Coronavirus cases by gender onboard the Princess Cruises ship (Female =313, Male = 321)

Out of the 634 confirmed cases, a total of 306 and 328 were reported to be symptomatic and asymptomatic, respectively. The proportion of asymptomatic individuals appears to be 16.1 % (35/218) before February 13, 25.6 % (73/285) on February 15, 31.2 % (111/355) on February 16, 39.9% (181/454) on February 17, 45.4 % (246/542) on February 18, 51.9% (322/621) on February 19 and 51.7% (328/634) on February 20 (Table 1). Soon after identification of the first infections, both symptomatic and asymptomatic cases were transported to designated medical facilities specialized in infectious diseases in Japan. However, these patients were treated as external (imported) cases, and a detailed description of their clinical progression is not publicly available.

**Table 1.** Test results for Passengers and crews of the Diamond Princess cruise (N = 3711)

| Date<sup>§</sup> | No of passenger and crew members onboard | No. of disembarked passenger and crew member (cumulative) | No. of test | No. of test (cumulative) | Positive | Positive (cumulative) | No. of symptomatic cases | No. of asymptomatic cases | No. of asymptomatic cases (cumulative) |
| --- | --- | --- | --- | --- | --- | --- | --- | --- | --- |
| Feb/5 | 3711 |  | 31 | 31 | 10 | 10 |  |  |  |
| Feb/6 |  |  | 71 | 102 | 10 | 20 |  |  |  |
| Feb/7 |  |  | 171 | 273 | 41 | 61 |  |  |  |
| Feb/8 |  |  | 6 | 279 | 3 | 64 |  |  |  |
| Feb/9 |  |  | 57 | 336 | 6 | 70 |  |  |  |
| Feb/10 |  |  | 103 | 439 | 65 | 135 |  |  |  |
| Feb/11 |  |  |  |  |  |  |  |  |  |
| Feb/12 |  |  | 53 | 492 | 39 | 174 |  |  |  |
| Feb/13 |  |  | 221 | 713 | 44 | 218 |  |  |  |
| Feb/14 | 3451 | 260<sup>‡</sup> |  |  |  |  |  |  |  |
| Feb/15 |  |  | 217 | 930 | 67 | 285 | 29 | 38 | 73<sup>¶</sup> |
| Feb/16 |  |  | 289 | 1219 | 70 | 355 | 32 | 38 | 111 |
| Feb/17 | 3183 | 528<sup>‡</sup> | 504 | 1723 | 99 | 454 | 29 | 70 | 181 |
| Feb/18 |  |  | 681 | 2404 | 88 | 542 | 23 | 65 | 246 |
| Feb/19 |  |  | 607 | 3011 | 79 | 621 | 11 | 68 | 322 |
| Feb/20 |  |  | 52 | 3063 | 13 | 634 | 7 | 6 | 328 |

<sup>§</sup>Reported date
<sup>‡</sup>As this is a cumulative number, the exact date of disembarkation is unavailable.
<sup>¶</sup>As this is a cumulative number, the reported date for 35 asymptomatic cases are unavailable.

The asymptomatic proportion was defined as the proportion of asymptomatically infected individuals among the total number of infected individuals.

### Statistical modelling

Here, we describe the statistical model that was employed to estimate the asymptomatic proportion using the time-series dataset described above.

The reported asymptomatic cases consists of both true asymptomatic infections and symptomatic cases that had not yet developed symptoms at the time of data collection, i.e., the data is right-censored. Each datum consists of an interval of time during which the individual may have been infected and a binary variable indicating whether they were symptomatic as of the 18th of February.

For individual i let [*a*_*i*,_ *b*_*i*_] denote the interval during which they may have been infected and *c* represents the censor date of observation of being symptomatic. The (unknown) time at which individual *i* was infected is denoted *X*_*i*_ and, if they develop symptoms, let *D*_*i*_ denote the delay from the time of infection until the time they are symptomatic, with cumulative density function (CDF), *F*_D_. The asymptomatic proportion, *p*, is the probability an individual will never develop symptoms.

Given an individual was exposed during the interval [*a*_*i*,_ *b*_*i*_] the probability for them being asymptomatic at time *c* is

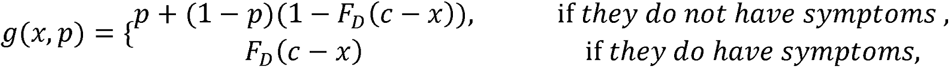

Given they were infected at time x for some *a*≤*x*≤*b*. Since the natural history of each individual’s infection is independent, the likelihood function is just the product of the *g*(*X*_i_, *p*) for each individual. Pervious work on COVID-19 suggests that the distribution of the delay, *D*, between infection and onset of symptomatic infections follows a Weibull distribution, with the mean and SD at 6.4 and 2.3 days [3]. The observations were treated as survival data with right-censoring. The probability of being asymptomatic along with the infection time of each individual where estimated in a Bayesian framework using Hamiltonian Monte Carlo (HMC). A detailed description of the model used and the computation is provided in a Technical Appendix.

## Findings from the real-time outbreak analysis

Posterior median estimates of true asymptomatic proportion among the reported asymptomatic cases is at 0.35 (95% CrI: 0.30–0.39), with the estimated total number of the true asymptomatic cases at 113.3 (95%CrI: 98.2-128.3) and the estimated asymptomatic proportion at 17.9% (95% CrI: 15.5%–20.2%).

We conducted sensitivity analyses to examine how varying the mean incubation period between 5.5 and 9.5 days affects our estimates of the true asymptomatic proportion. Estimates of the true asymptomatic proportion among the reported asymptomatic cases are somewhat sensitive to changes in the mean incubation period, ranging from 0.28 (95%CrI: 0.23–0.33) to 0.40 (95%CrI: 0.36–0.44), while the estimated total number of true asymptomatic cases range from 91.9 (95%CrI: 75.2–108.7) to 130.8 (95%CrI: 117.1–144.5) and the estimated asymptomatic proportion ranges from 20.6% (95%CrI: 18.5%–22.8%) to 39.9% (95%CrI: 35.7%–44.1%).

Heat maps were used to display the density distribution of infection timing by individuals (Figure S1) where the vertical line corresponds to the date of February 5, 2020. Among the symptomatic cases, the infection timing appears to have occurred just before or around the start of the quarantine period, while the infection timing for asymptomatic cases appears to have occurred well before the start of the quarantine period.

## Discussion

We have conducted statistical modeling analyses on publicly available data to elucidate the asymptomatic proportion, along with the time of infection among the COVID-19 infected cases onboard the Princess Cruises ship.

Our estimated asymptomatic proportion is at 17.9% (95% CrI: 15.5%–20.2%), which overlaps with a recently derived estimate of 33.3% (95% CI: 8.3%–58.3%) from data of Japanese citizens evacuated from Wuhan [13]. Considering the similarity in viral loads and the high possibility of potent transmission potential, the high proportion of asymptomatic infections has significant public health implications [14]. For instance, self-isolation for 14-day periods are also recommended for contacts with asymptomatic cases [15].

Most of the infections appear to have occurred before or around the start of the 2-week quarantine that started on February 5, 2020, which further highlights the potent transmissibility of the SARS-CoV-2 virus, especially in confined settings. To further mitigate transmission of COVID-19 and bring the epidemic under control in areas with active transmission, it may be necessary to minimize the number of gatherings in confined settings.

Our study is not free from limitations. First, laboratory tests by PCR were conducted focusing on symptomatic cases especially at the early phase of the quarantine. If asymptomatic cases where missed as a result of this, it would mean we have underestimated the asymptomatic proportion. Second, it is worth noting that the data of passengers and crews employed in our analysis is not a random sample from the general population. Considering that most of the passengers are 60 years and older, the nature of this age distribution may lead to underestimation if older individuals tend to experience more symptoms. An age standardized asymptomatic proportion would be more appropriate in that case. Third, the presence of symptoms in cases with COVID-19 may correlate with other factors unrelated to age including prior health conditions such as cardiovascular disease, diabetes, immunosuppression. Therefore, more detailed data documenting the baseline health of the individuals including the presence of underlying diseases or comorbidities would be useful to remove the bias in estimates of the asymptomatic proportion.

In summary, we have estimated the proportion of asymptomatic cases among individuals who have tested positive for novel COVID-19 along with the times of infection of confirmed cases onboard the Princess Cruises Ship after adjusting for the delay in symptom onset and right-censoring of the observations.

## Data Availability

The present study relies on published data and access information to essential components of the data are available from the corresponding author.

## Additional files

### Additional file 1: Supplementary document

#### Additional file 2: Figure S1. Heat maps of the density distribution of infection timing by individuals

A) Symptomatic cases (N= 306), B) Asymptomatic cases (N= 328). Vertical axis represents each individual from 1 to N. Cases disembarked after testing positive for the disease. Day 1 corresponds to January 20, 2020, when the first symptomatic case embarked. The vertical line corresponds to February 5, 2020 when the quarantine period started.

## REFERENCES

1. World Health Organization (WHO). Novel Coronavirus (2019-nCoV) situation reports. Available from: https://www.who.int/emergencies/diseases/novel-coronavirus-2019/situation-reports

2. Linton NM, Kobayashi T, Yang Y, Hayashi K, Akhmetzhanov AR, Jung SM, et al. Epidemiological characteristics of novel coronavirus infection: A statistical analysis of publicly available case data. medRxiv 2020.01.26.20018754; doi: https://doi.org/10.1101/2020.01.26.20018754

3. Backer JA, Klinkenberg D, Wallinga J. Incubation period of 2019 novel coronavirus (2019-nCoV) infections among travelers from Wuhan, China, 20-28 January 2020. Euro Surveill. 2020;25(5). Doi: 10.2807/1560-7917.ES.2020.25.5.2000062.

4. Kroon FP, Weiland HT, van Loon AM, van Furth R. Abortive and subclinical poliomyelitis in a family during the 1992 epidemic in The Netherlands. Clin Infect Dis. 1995;20:454–456.

5. Mbabazi WB, Nanyunja M, Makumbi I, et al. Achieving measles control: lessons from the 2002-06 measles control strategy for Uganda. Health Policy Plan. 2009;24:261–269.

6. Smallman-Raynor MR, Cliff AD, eds. Poliomyelitis: Emergence to Eradication. New York: Oxford University Press; 2006;32.

7. Miura F, Matsuyama R, Nishiura H. Estimating the Asymptomatic Ratio of Norovirus Infection During Foodborne Outbreaks With Laboratory Testing in Japan.J Epidemiol. 2018;28(9):382–387. Doi: 10.2188/jea.JE20170040.

8. Mizumoto K, Kobayashi T, Chowell G. Transmission potential of modified measles during an outbreak, Japan, March□May 2018. Euro Surveill. 2018 Jun;23(24). Doi: 10.2807/1560-7917.ES.2018.23.24.1800239.

9. The Novel Coronavirus Pneumonia Emergency Response Epidemiology Team. The Epidemiological Characteristics of an Outbreak of 2019 Novel Coronavirus Diseases (COVID-19) — China, 2020[J]. China CDC Weekly 2020.

10. HoehlS, Berger A, Kortenbusch M, Cinatl J, Bojkova D, Rabenau H, et al. Evidence of SARS-CoV-2 Infection in Returning Travelers from Wuhan, China. N Engl J Med. 2020. Doi: 10.1056/NEJMc2001899.

11. Huang C, Wang Y, Li X, et al.Clinical features of patients infected with 2019 novel coronavirus in Wuhan, China. Lancet. 2020. Pii: S0140-6736(20)30183-5.

12. Minister of Health, Labour and Welfare, Japan. Available from https://www.mhlw.go.jp/stf/seisakunitsuite/bunya/0000164708_00001.html [In Japanese]

13. Nishiura H, Kobayashi T, Miyama T, Suzuki A, Jung S, Hayashi K,et al. Estimation of the asymptomatic ratio of novel coronavirus infections (COVID-19) doi: https://doi.org/10.1101/2020.02.03.20020248

14. Zou L, Ruan F, Huang M, Liang L, Huang H, Hong Z, et al. SARS-CoV-2 Viral Load in Upper Respiratory Specimens of Infected Patients. N Engl J Med. 2020. Doi: 10.1056/NEJMc2001737

15. Center for DIsease Control and Prevention, USA. Interim US Guidance for Risk Assessment and Public Health Management of Persons with Potential Coronavirus Disease 2019 (COVID-19) Exposure in Travel-associated or Community Settings.Updated February 8, 2020. Available from https://www.cdc.gov/coronavirus/2019-ncov/php/risk-assessment.html [accessed March 2, 2020]

